# Equalizing prognostic disparities in stage III KRAS-mutant NSCLC: addition of durvalumab to combined chemoradiotherapy improves survival

**DOI:** 10.1101/2024.03.21.24304669

**Authors:** Ella A. Eklund, Mathilda Orgard, Delice Wallin, Sama I. Sayin, Henrik Fagman, Sukanya Raghavan, Levent M Akyürek, Jan Nyman, Clotilde Wiel, Andreas Hallqvist, Volkan I. Sayin

## Abstract

**Introduction:** Stage III non-small cell lung cancer (NSCLC) is heterogeneous and identification of subgroups with differential responses is crucial to optimize treatment. Addition of durvalumab to concurrent chemoradiotherapy (cCRT) has previously been shown to improve survival outcomes. Meanwhile, subgroups harboring KRAS mutations have been shown to have worse prognosis. We investigated whether KRAS mutational status may affect survival outcomes after adjuvant durvalumab following cCRT in stage III NSCLC.

**Methods:** In this retrospective study, we present a real-world dataset of all stage III NSCLC patients treated with cCRT with a curative intent and molecularly assessed between 2016-2021 in West Sweden. Primary study outcomes were overall survival (OS) and progression free survival (PFS).

**Results:** We identified 145 patients receiving cCRT with a curative intent, 32% harbored an activating mutation in the KRAS gene (KRAS^MUT^). Compared to KRAS wild-type (KRAS^WT^), KRAS^MUT^ had a worse OS (*p*=0.047) and PFS (*p*=0.038). The finding persisted on multivariate analysis with OS (HR 1.703, 95%CI 1.074-2.702, *p* = 0.024) and PFS (HR 1.628, 95% CI 1.081-2.453, *p* = 0.020). After the addition of durvalumab to cCRT, there were no longer any significant differences between KRAS^WT^ and KRAS^MUT^ in OS or PFS.

**Conclusions:** *KRAS* mutations are a negative prognostic factor after cCRT in stage III NSCLC, and the addition of durvalumab equalizes the negative impact of harboring this mutation.

## Introduction

Lung cancer remains the leading cause of cancer mortality, and non-small cell lung cancer (NSCLC) accounts for 80-85% of cases. Stage at diagnosis is an important prognostic factor in NSCLC, since patients in later stages have increasingly worse prognosis.^1^ Staging (I-IV) is based on the TNM system, which describes the size and infiltration status of the primary tumor, number and location of regional lymph nodes involved, and number and location of metastases. About 30% of new cases of NSCLC are stage III at diagnosis. Stage III, also referred to as locally advanced NSCLC, is defined as having spread locoregionally through primary tumor extension into extrapulmonary structures and involving hilar or mediastinal lymph nodes but having no evidence of distant metastases.^2-4^

Importantly, stage III NSCLC is a heterogenous group, with overall survival (OS) rates ranging between 35% and 10%.^2, 5, 6^ There are large variations between patients belonging to this group in the degree of local disease advancement, clinical presentation, and treatment options.^7-9^ Treatment regimens are generally determined in multidisciplinary case discussions at high volume centers including oncologists, radiologists, pneumologists and thoracic surgeons.^10, 11^ Treatment of stage III NSCLC patients can be highly personalized and the identification of subgroups that are more responsive to certain treatment combinations than others is

In recent years there has been growing interest in KRAS-mutated (KRAS^MUT^) NSCLC due to the introduction of new tyrosine kinase inhibitors (TKI) targeting KRAS, offering new treatment possibilities for this subgroup. KRAS mutation is the most prevalent oncogenic driver in NSCLC (35%) but the prognostic and predictive role of KRAS mutations is under debate. While we and others have shown KRAS mutations to negatively influence the prognosis of NSCLC,^12-15^ some studies found no difference in OS.^16-18^ In addition, some have reported shorter OS for KRAS^MUT^ compared to wild type (KRAS^WT^) patients following first-line platinum-based chemotherapy.^19-21^ Hence, the significance of KRAS mutation for prognosis and treatment response in NSCLC disease remains a pending topic.^22^

First-line treatment for unresectable stage III NSCLC is concurrent chemoradiotherapy (cCRT) provided that patients have a decent performance status, adequate lung function and no discouraging comorbidity. Treatment is typically given with a platinum doublet and conventionally fractioned radiotherapy to a total dose of 60-66 Gy.

Addition of immune check point blockades were first approved in stage IV treatment schedules and was found to be associated with substantial improvements in overall survival, marking a new paradigm in lung cancer care.^13, 14^ Patients in this group receive targeted therapy depending on specific mutations if present, or immunotherapy-containing regimens, either as monotherapy or in combination with chemotherapy or a second immunotherapy drug.^23^ Consolidation therapy with the programmed death-ligand (PD-L1) inhibitor durvalumab after chemoradiotherapy in stage III NSCLC was approved in 2018 Europe based on findings from the PACIFIC trial.^15^ This trial demonstrated the superiority of durvalumab over placebo in terms of increased overall survival among patients with stage III NSCLC treated with chemoradiotherapy. Several studies reporting similar findings have followed ^16^, and recently a 5-year follow up of the PACIFIC trial showed that these changes persisted over time.^17^ Durvalumab is now part of standard treatment regimens for stage III NSCLC patients where those with no disease progression after first-line therapy with cCRT and PD-L1 expression ≥ 1 %, are recommended adjuvant treatment with durvalumab for 12 months.^18^

Although relatively few studies have been conducted regarding the impact of KRAS mutation on prognosis after cCRT in stage III NSCLC, the available data suggests KRAS mutation as a negative predictor for treatment response and OS.^20, 21^ Furthermore, data on KRAS^MUT^ in the context of ICB treatment in stage III disease is scarce. In summary, given previous findings of heterogeneity of stage III NSCLC, worsened prognosis in the presence of KRAS mutations in this group, and improved prognosis after the introduction of consolidation therapy with durvalumab in the overall group, we wondered whether KRAS mutation subgroups may respond differentially to durvalumab. Here, we investigate whether consolidation therapy with durvalumab affects survival outcomes in KRAS mutated stage III NSCLC patients. Our aim with this study is to assess outcomes in KRAS^MUT^ vs KRAS^WT^ in locally advanced NSCLC patients receiving cCRT with and without durvalumab.

## Material and Methods

### Study Design and Study Population

This retrospective cohort study includes all consecutive patients with stage III NSCLC in the Västra Götaland region, Sweden, between the years 2016-2021 who underwent molecular assessment and received combined chemoradiotherapy with curative intent (*n* = 145). Patients were excluded if they underwent surgery or received sequential chemoradiotherapy (*n* = 45) (Figure 1). The study period encompasses the standard practice before the introduction of durvalumab, allowing for the evaluation of the potential impact of ICB. For inclusion in the durvalumab analyses, a single dose of durvalumab was deemed sufficient.

**Figure 1.**
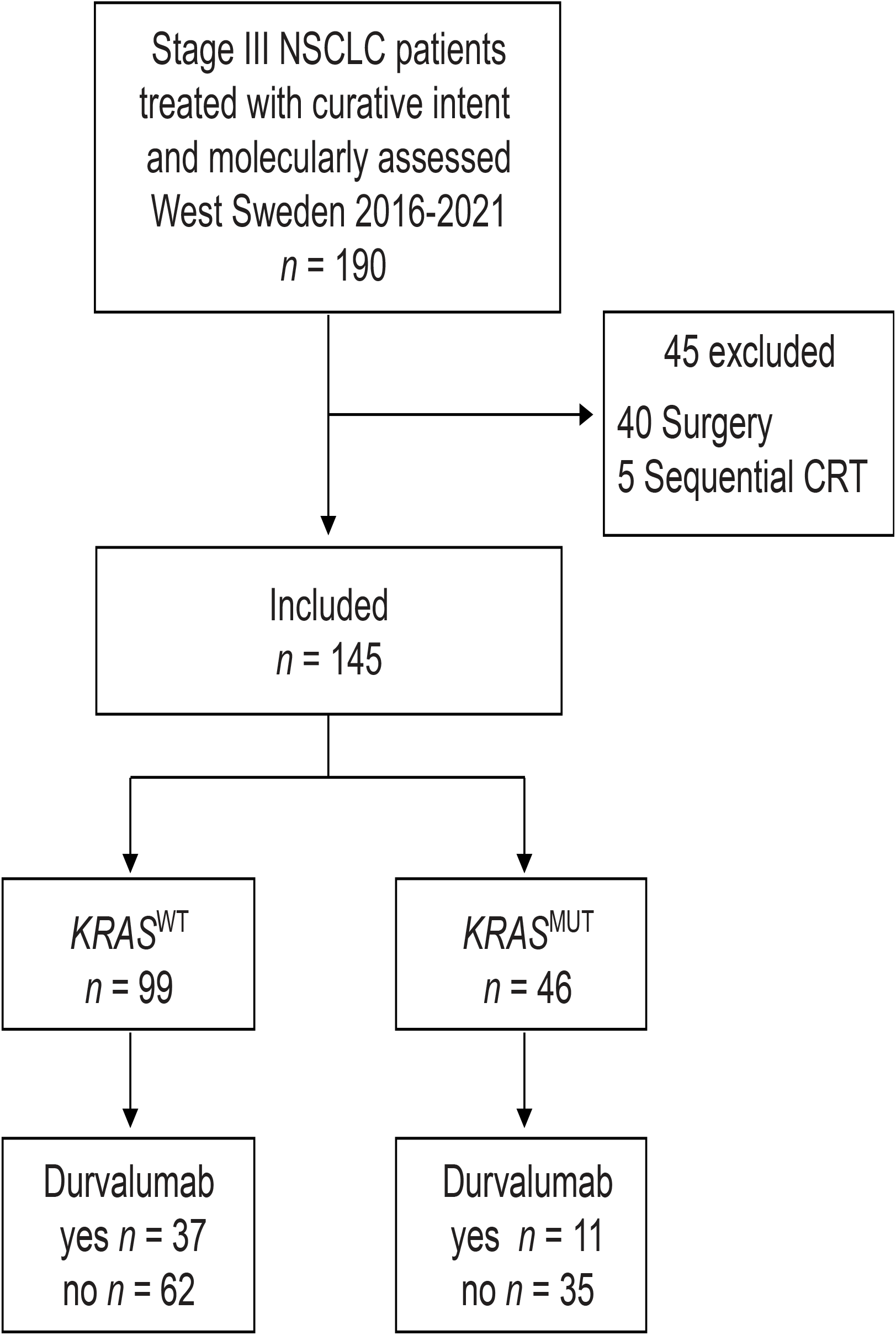
Patient selection. Flow chart showing patient selection for the study. NSCLC: Non-small cell lung cancer; CRT: chemoradiotherapy; *KRAS*^WT^: *KRAS* wildtype; *KRAS*^MUT^: *KRAS* mutated

### Data Collection Procedures

Patients were selected for the study through the pathology regional laboratory information system, where they were referred for molecular assessment. The selected patients underwent next-generation sequencing (NGS) on DNA from FFPE blocks or cytological smears using the Ion AmpliSeq™ Colon and Lung Cancer Panel v2 from Thermo Fisher Scientific until 2019 and thereafter the Thermo Fisher Oncomine™ Focus Assay, assessing hotspot mutations in *EGFR, BRAF, KRAS* and *NRAS*. Until June 2017, *ALK*-fusions were assessed with immunohistochemistry (IHC), and with fluorescence in situ hybridization (FISH) if positive or inconclusive IHC; ROS1 was analyzed upon request with FISH. Thereafter, *ALK, ROS1* and *RET* fusions were assessed on RNA using the Oncomine Solid Tumor Fusion Panel from Thermo Fisher Scientific. The analyzes was done as a part of the diagnostic workup process at the Department of Clinical Pathology at Sahlgrenska University Hospital

Demographic data and pathological details were retrieved from the Swedish Lung Cancer Registry and patient charts. The trial was approved by the Swedish ethical review authority (Dnr 2019-04771, 2021-04987) and conducted in accordance with the Declaration of Helsinki and Good Clinical Practice guidelines as defined by the International Conference on Harmonization.

### Variables

Collected patient demographic details included age, sex, Eastern Cooperative Oncology Group performance status (ECOG PS) and smoking history (current/former/never).

Pathological details encompassed substage, histology and mutational status. All patients were classified according TNM 8^th^ edition. Treatment characteristics included platinum-doublet chemotherapy regimen, total radiation dose and administration of durvalumab. Time-to-event data included the date of treatment initiation, the date of disease progression and/or death and the date of last follow-up.

### Outcomes

The primary endpoints were overall survival (OS) and progression free survival (PFS). OS was defined as the time interval between the date of first treatment and the date of death from any cause. PFS was defined as the time interval between the date of first treatment and the date of progression or death whichever came first. Patients without disease progression and were still alive at the time of data collection were censored at last follow-up. OS and PFS, stratified based on KRAS^MUT^ versus KRAS^WT^ patients, were analyzed both for the entire cohort and subgroups receiving or not receiving durvalumab.

### Statistical Analysis

Descriptive statistics were used to describe the clinical characteristics, which were further evaluated using univariate analysis. OS and PFS, stratified by KRAS mutational status, PDL1 and treatment type, were estimated using the Kaplan-Meier method. Log-rank tests were used to determine significant differences in PFS and OS between groups. Multivariable Cox regression analysis was conducted to compensate for potential confounders. Median follow-up time was calculated using the reverse Kaplan-Meier method. Statistical significance was set at p < 0.05.

## Results

### Patients and tumor characteristics

A total number of 145 patients were included in the study (Figure 1) and their characteristics are shown in Table 1. The median age at diagnosis was 69 years for both KRAS^WT^ patients and KRAS^MUT^ patients. There were more females and a more substantial smoking history in the KRAS^MUT^ population. ECOG PS 0 was more common among KRAS^MUT^ (45%) than KRAS^WT^ patients (34%). A greater proportion of KRAS^WT^ patients had substage IIIC at diagnosis. Squamous cell carcinoma was non-existent in the KRAS^MUT^ group but common in the KRAS^WT^ group (40%). The majority of KRAS^MUT^ patients had an adenocarcinoma (89.%). The most common driver mutations were in KRAS and EGFR. KRAS^G12C^ was the most common KRAS sub-mutation (24, 52%). Median follow-up time was 48 months (95% CI 40-48).

### KRAS mutation worsens survival outcomes in stage III NSCLC patients

Among our entire study population (n=145) there was a significantly lower median OS among KRAS^MUT^ patients (25 months) compared with KRAS^WT^ (46 months; *p*=0.047). Additionally, PFS was significantly lower among KRAS^MUT^ patients (9 months) compared to 16 months for KRAS^WT^ (16 months; *p*= 0.038) (Figure 2A & B). The results were further controlled for confounders with multivariate cox regression. We found that KRAS mutation was an independent prognostic factor for worse OS (HR 1.703, 95% CI 1.074-2.702, *p* = 0.024) and PFS (HR 1.628, 95% CI 1.081-2.453, *p* = 0.020) (Figure 2C & D).

**Figure 2.**
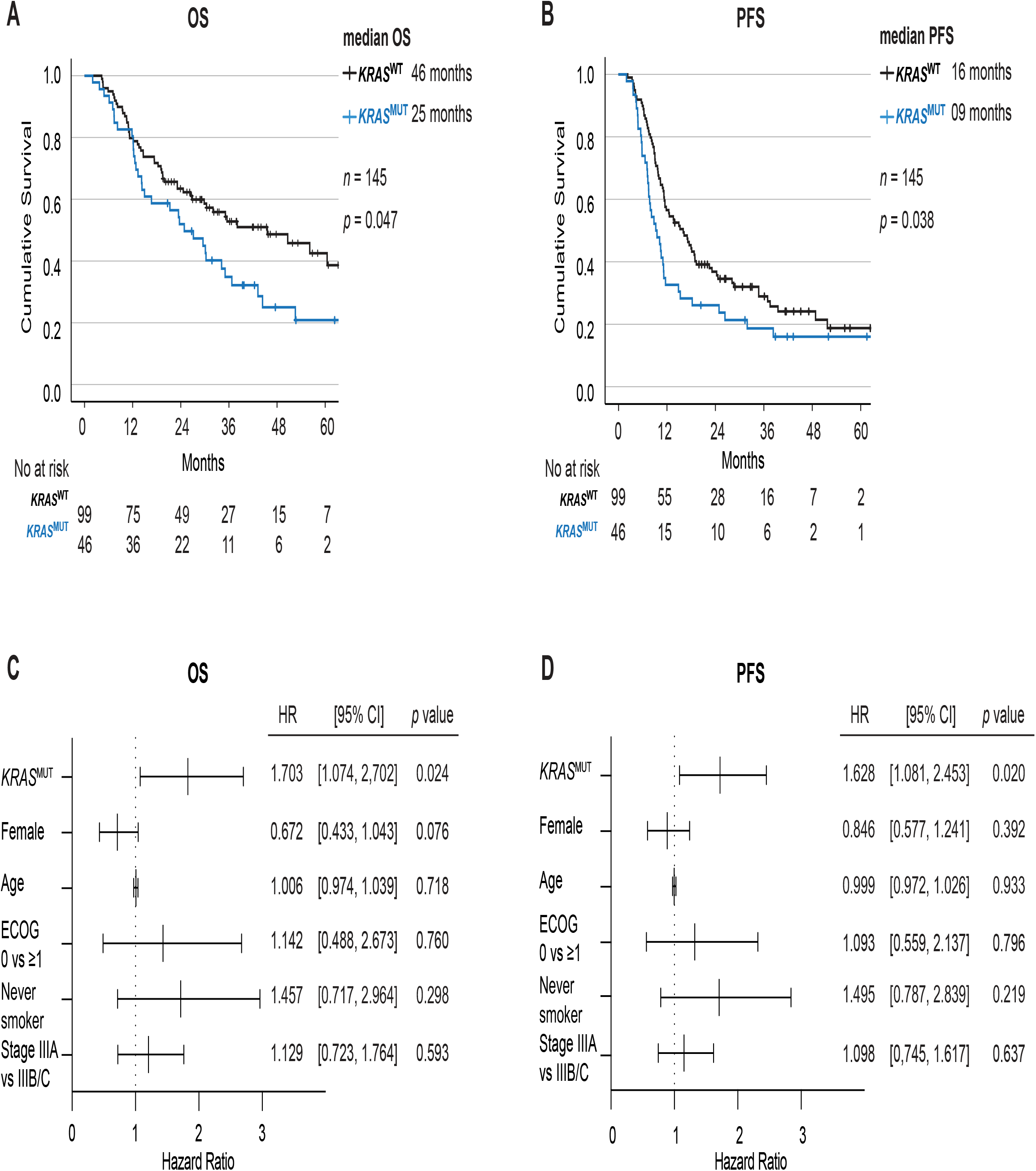
KRAS mutation worsens survival outcomes in stage III NSCLC patients. Kaplan-Meier estimates comparing overall survival (**A)** and progression free survival (**B)** stratified on KRAS status for all patients in the cohort. Forest plot of multivariate COX regression analysis for overall survival (**C**) and progression free survival (**D**). OS: overall survival; PFS: progression free survival; NR Not reached, cCRT: concurrent chemoradiotherapy; HR: Hazard Ratio; CI: Confidence of interval; *KRAS*^WT^: *KRAS* wildtype; *KRAS*^MUT^: *KRAS* mutated

### KRAS mutation worsens survival outcomes after cCRT among stage III NSCLC patients

When studying the impact of KRAS mutational status on prognosis following cCRT, we found significantly worse median OS for KRAS^MUT^ (24 months) vs KRAS^WT^ (35 months) (*p* = 0.036) (Figure 3A) and, additionally, worse median PFS of 8 vs 13 months (*p* = 0.037), respectively (Figure 3B).

**Figure 3.**
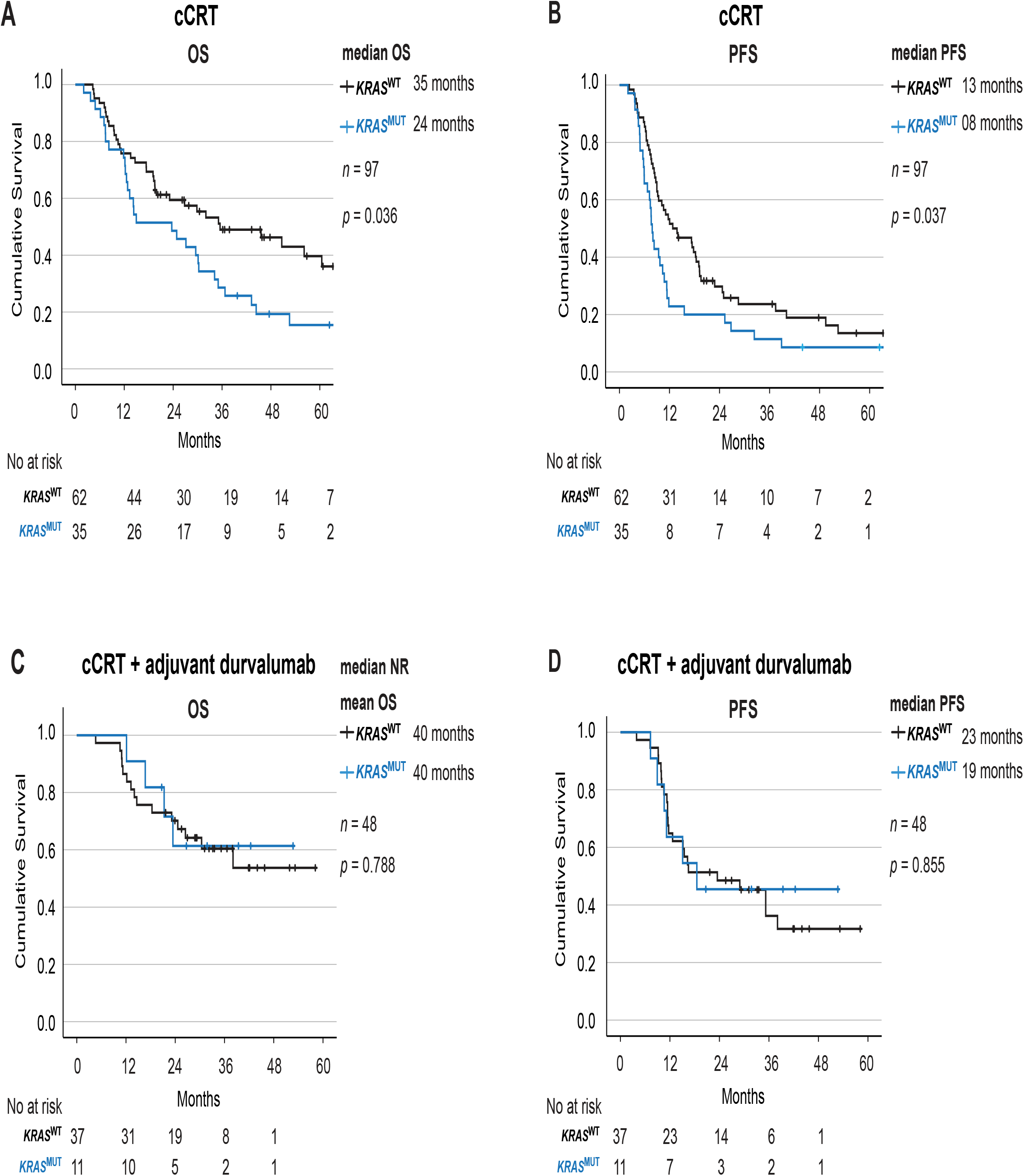
Adjuvant durvalumab equalizes the negative impact of KRAS mutation on survival outcomes after cCRT. Kaplan-Meier estimates comparing overall survival (**A)** and progression free survival (**B)** stratified on KRAS status for all patients receiving cCRT. Kaplan-Meier estimates comparing overall survival (**C)** and progression free survival (**D)** stratified on KRAS status for all patients receiving cCRT and adjuvant durvalumab. OS: overall survival; PFS: progression free survival; NR Not reached, cCRT: concurrent chemoradiotherapy; *KRAS*^WT^: *KRAS* wildtype; *KRAS*^MUT^: *KRAS* mutated

### Adjuvant durvalumab significantly improves survival outcomes after cCRT among all stage III NSCLC patients

When analyzing the effect of consolidation durvalumab on the full cohort we observed a significant improved OS and PFS with median OS not reached for the durvalumab group (mean OS 41 months) and 30 months for the group not receiving durvalumab (*p* = 0.037) and a median PFS of 19 months vs 10 months, respectively (*p* < 0.001)(Supplemental figure 1A & B).

### KRAS mutation groups show improved survival outcomes after adjuvant durvalumab

While KRAS^MUT^ groups showed worse prognosis (24 months) than wildtype (35 months) after treatment with cCRT alone (*p* = 0.036) (Figure 3A), addition of durvalumab to the treatment regimen equalized this difference and KRAS^MUT^ patients had no longer any significant differences in survival outcomes (40 months) from KRAS^WT^ (40 months) (*p* = 0.788) (Figure 3C). Similarly, when measuring PFS, while KRAS^MUT^ had significantly decreased PFS (8 months) compared to KRAS^WT^ (13 months) among patients treated with cCRT alone (p = 0.037) (Figure 3B), addition of durvalumab to the treatment regimen abolished the additionally negative prognosis of KRAS^MUT^, and this group had no longer had significant difference in PFS (19 months) compared to KRAS^WT^ (23 months) (*p* = 0.855) (Figure 3D).

### Adjuvant durvalumab equalizes the negative impact of KRAS mutation on survival outcomes after cCRT

To further investigate whether KRAS^MUT^ is a subgroup in stage III NSCLC that especially gains from adjuvant therapy with durvalumab after cCRT, we analyzed the effect of treatment within each mutational group separately. We observed that treatment with durvalumab had greater effect on improving OS in KRAS^MUT^ group (*p* = 0.063) compared to KRAS^WT^ (*p* = 0.345) (Supplemental figure 2A, 2C). Both KRAS^WT^ (*p* = 0.033) and KRAS^MUT^ (*p* = 0.011) groups had a significant benefit of adjuvant durvalumab on PFS (Supplemental figure 2B, 2D).

## Discussion

This study suggests that the previously inferior prognosis of stage III KRAS^MUT^ compared to KRAS^WT^ patients treated with cCRT has improved to a similar outcome in both groups due to the introduction of ICB in the stage III setting. There is no survival difference between the KRAS populations when durvalumab is administered in contrast to the findings from the era before ICB. The predictive role of KRAS mutation in relation to outcome in NSCLC is a controversial topic, particularly for stage III NSCLC where the literature is relatively sparse in contrast to earlier stages and metastatic stage IV disease. In the latter, treated with chemotherapy, some studies describe KRAS mutation as a negative prognostic factor for OS and PFS ^13-15^ while others demonstrate an equal survival benefit for KRAS^MUT^ and KRAS^WT^ patients^16-18^. After the introduction of ICB in the stage IV treatment paradigm new assessment of the impact of KRAS have been performed but do not consistently point in the same direction, and the predictive value of KRAS related to ICB treatment remains debatable. ^21^ The prognostic value of mutated KRAS in stage III patients treated with concurrent chemoradiotherapy likely indicates an association with an inferior prognosis, as suggested by previous data where mutated KRAS was independently connected to a decrease in overall survival in multivariate analysis.^22, 24^ Regarding the significance of KRAS mutations in stage III disease after the introduction of durvalumab, there are a few reported cohorts. Riudavets et al observed in a study with 323 patients that there was no PFS difference between patients with a mix of oncogenic drivers compared to wild type patients^25^. In the group with oncogenic drivers (n=43) the KRAS^G12C^ mutated stage III NSCLC patients had superior PFS compared to other driver genetic alterations (EGFR^del119/ex21^, BRAF^V600E^, ALK-rearrangements) following consolidative durvalumab treatment but no difference in OS was seen at a median follow-up time of 18 months. This finding is in line with previous data where oncogenic drivers usually associated with never smokers are known not to correlate with response to ICB treatment,^26^ at least not to ICB monotherapy.^27^ Liu et al observed in a cohort of 104 patients a shorter PFS for both KRAS^MUT^ and non-KRAS driver variations compared to KRAS^WT^ in stage III NSCLC and no difference in OS was found at a median follow-up time of 23.6 months.^28^ In accordance with the current study, Guo et al recognized KRAS mutated stage III NSCLC as having equal PFS and OS as tumors with no driver mutations in a cohort of 74 patients.^8^ Lastly, in a recent study by Cortiula et al they assessed outcome in a cohort of 66 patients harbouring KRAS mutations and uncommon genetic alterations (PIK3CA, TP53, MET, BRAF, HER2, uncommon EGFR) where a beneficial effect of durvalumab on OS and PFS was demonstrated.^29^

Another potential improvement strategy is the addition of KRAS inhibitors in the stage III treatment schedule. At present sotorasib is the only approved drug, but only in stage IV NSCLC second line.^30^ In theory TKI’s could be added to the stage III approach, either up-front, or as an adjuvant therapy in line with the addition of EGFR and ALK-directed therapy after surgery.^31, 32^

The findings herein are limited by the retrospective design and low sample size, furthermore the impact of co-mutational partners like KEAP1 or STK11 is not known. Nevertheless, the results represent a consecutive cohort in a real-world setting. The findings contribute the knowledge in line with other small cohorts and in conclusion: addition of durvalumab after cCRT in stage III NSCLC patients has improved outcome for the KRAS^MUT^ patients to a similar level as for the KRAS^WT^ group.

## Acknowledgements

We thank members of the Swedish Lung Cancer Registry and the continuous reporting by Swedish healthcare employees. This work was supported by the Swedish Research Council (2018-02318 and 2022-00971 to VIS, 2021-03138 to CW), the Swedish Cancer Society (23-3062 to VIS, 22-0612FE to CW), the Gothenburg Society of Medicine (2019; 19/889991 to EAE), Assar Gabrielsson Research Foundation (to EAE, CW, and VIS), the Swedish state under the agreement between the Swedish government and the county councils, the ALF-agreement (to HF), Department of Oncology, Sahlgrenska University Hospital (to EAE and AH), the Swedish Society for Medical Research (2018; S18-034 to VIS), the Knut and Alice Wallenberg Foundation, and the Wallenberg Centre for Molecular and Translational Medicine (to VIS).

## Authors’ contributions

Conceptualization, E.A.E., A.H. and V.I.S.; Data curation, E.A.E., M.O., D.W., H.F.; Formal analysis, E.A.E., M.O. and D.W.; Funding acquisition, E.A.E., H.F., S.R., L.M.A., J.N., C.W., A.H. and V.I.S.; Methodology, E.A.E.; Resources, H.F.; Supervision, A.H. and V.I.S.; Visualization, E.A.E., S.I.S., C.W. and V.I.S.; Writing—original draft, E.A.E., S.I.S, A.H. and V.I.S.; Writing—review & editing, E.A.E., S.I.S., H.F., S.R., L.M.A., J.N., C.W., A.H. and V.I.S., Project coordination, A.H. and V.I.S. All authors have read and agreed to the published version of the manuscript.

## Institutional Review Board Statement

Approval from the Swedish Ethical Review Authority (Dnr 2019-04771 and 2021-04987) was obtained prior to the commencement of the study. No informed consent was required due to all data presented in a de-identified form according to the Swedish Ethical Review Authority.

## Consent for publication

Not applicable. Patient consent statements were not required due to the retrospective nature of this study. No informed consent was required due to all data presented in a de-identified form according to the Swedish Ethical Review Authority.

## Data Availability Statement

The datasets used and/or analyzed during the current study available from the corresponding author on reasonable request.

## List of Supplemental material

Supplemental Figure 1. PDF

Supplemental Figure 2. PDF

## Figures legend

**Table 1** Characteristics of the total cohort as well as stratified by *KRAS*^WT^ and *KRAS*^MUT^. Data are presented as n (%).

cCRT: concurrent chemoradiotherapy; *KRAS*^WT^: *KRAS* wildtype; *KRAS*^MUT^: *KRAS* mutated; ECOG PS, Eastern Cooperative Oncology Group Performance Status; NSCLC NOS: non-small cell lung cancer not otherwise specified; Gy: Gray

**Supplemental Figure 1 Adjuvant durvalumab significantly improves survival outcomes after cCRT among all stage III NSCLC patients**

Kaplan-Meier estimates comparing overall survival (**A)** and progression free survival (**B)** stratified on cCRT +/- adjuvant durvalumab.

OS: overall survival; PFS: progression free survival; cCRT: concurrent chemoradiotherapy

**Supplemental Figure 2 Improved survival outcomes after adjuvant durvalumab and cCRT among stage III NSCLC patients harboring KRAS mutations**

Kaplan-Meier estimates comparing overall survival (**A)** and progression free survival (**B)** stratified on cCRT +/- adjuvant durvalumab for all KRAS wild type patients. Kaplan-Meier estimates comparing overall survival (**C)** and progression free survival (**D)** Kaplan-Meier estimates comparing overall survival (**A)** and progression free survival (**B)** stratified on cCRT +/- adjuvant durvalumab for all KRAS mutated patients.

OS: overall survival; PFS: progression free survival; NR Not reached, cCRT: concurrent chemoradiotherapy; *KRAS*^WT^: *KRAS* wildtype; *KRAS*^MUT^: *KRAS* mutated

## References

1. Sung H, Ferlay J, Siegel RL, et al. Global Cancer Statistics 2020: GLOBOCAN Estimates of Incidence and Mortality Worldwide for 36 Cancers in 185 Countries. CA Cancer J Clin 2021;71:209–249.

2. Lungcancer Nationellt Vårdprogram Version 6.0. Regionala Cancercentrum i Samverkan: 2022-05-10.

3. Goldstraw P, Chansky K, Crowley J, et al. The IASLC Lung Cancer Staging Project: Proposals for Revision of the TNM Stage Groupings in the Forthcoming (Eighth) Edition of the TNM Classification for Lung Cancer. J Thorac Oncol 2016;11:39–51.

4. König D, Savic Prince S, Rothschild SI. Targeted Therapy in Advanced and Metastatic Non-Small Cell Lung Cancer. An Update on Treatment of the Most Important Actionable Oncogenic Driver Alterations. Cancers 2021;13.

5. Majem M, Hernandez-Hernandez J, Hernando-Trancho F, et al. Multidisciplinary consensus statement on the clinical management of patients with stage III non-small cell lung cancer. Clin Transl Oncol 2020;22:21–36.

6. Casal-Mourino A, Ruano-Ravina A, Lorenzo-Gonzalez M, et al. Epidemiology of stage III lung cancer: frequency, diagnostic characteristics, and survival. Transl Lung Cancer Res 2021;10:506–518.

7. Ferrer I, Zugazagoitia J, Herbertz S, et al. KRAS-Mutant non-small cell lung cancer: From biology to therapy. Lung Cancer 2018;124:53–64.

8. Guo MZ, Murray JC, Ghanem P, et al. Definitive Chemoradiation and Durvalumab Consolidation for Locally Advanced, Unresectable KRAS-mutated Non-Small Cell Lung Cancer. Clin Lung Cancer 2022;23:620–629.

9. Mascaux C, Iannino N, Martin B, et al. The role of RAS oncogene in survival of patients with lung cancer: a systematic review of the literature with meta-analysis. Br J Cancer 2005;92:131–139.

10. Johnson ML, Sima CS, Chaft J, et al. Association of KRAS and EGFR mutations with survival in patients with advanced lung adenocarcinomas. Cancer 2013;119:356–362.

11. Guan JL, Zhong WZ, An SJ, et al. KRAS mutation in patients with lung cancer: a predictor for poor prognosis but not for EGFR-TKIs or chemotherapy. Ann Surg Oncol 2013;20:1381–1388.

12. Meng D, Yuan M, Li X, et al. Prognostic value of K-RAS mutations in patients with non-small cell lung cancer: a systematic review with meta-analysis. Lung Cancer 2013;81:1–10.

13. Marabese M, Ganzinelli M, Garassino MC, et al. KRAS mutations affect prognosis of non-small-cell lung cancer patients treated with first-line platinum containing chemotherapy. Oncotarget 2015;6:34014–34022.

14. Hames ML, Chen H, Iams W, et al. Correlation between KRAS mutation status and response to chemotherapy in patients with advanced non-small cell lung cancerZl. Lung Cancer 2016;92:29–34.

15. Eklund EA, Wiel C, Fagman H, et al. KRAS Mutations Impact Clinical Outcome in Metastatic Non-Small Cell Lung Cancer. Cancers (Basel) 2022;14.

16. Brady AK, McNeill JD, Judy B, et al. Survival outcome according to KRAS mutation status in newly diagnosed patients with stage IV non-small cell lung cancer treated with platinum doublet chemotherapy. Oncotarget 2015;6:30287–30294.

17. Mellema WW, Dingemans AM, Thunnissen E, et al. KRAS mutations in advanced nonsquamous non-small-cell lung cancer patients treated with first-line platinum-based chemotherapy have no predictive value. J Thorac Oncol 2013;8:1190–1195.

18. Rodenhuis S, Boerrigter L, Top B, et al. Mutational activation of the K-ras oncogene and the effect of chemotherapy in advanced adenocarcinoma of the lung: a prospective study. J Clin Oncol 1997;15:285–291.

19. Goulding RE, Chenoweth M, Carter GC, et al. KRAS mutation as a prognostic factor and predictive factor in advanced/metastatic non-small cell lung cancer: A systematic literature review and meta-analysis. Cancer Treat Res Commun 2020;24:100200.

20. Ma SX, Ma N, Han J, et al. [The efficacy and prognostic factors of immunotherapy in advanced non-small cell lung cancer patients with different driver gene mutations]. Zhonghua Yi Xue Za Zhi 2022;102:922–929.

21. Uehara Y, Watanabe K, Hakozaki T, et al. Efficacy of first-line immune checkpoint inhibitors in patients with advanced NSCLC with KRAS, MET, FGFR, RET, BRAF, and HER2 alterations. Thorac Cancer 2022;13:1703–1711.

22. Yagishita S, Horinouchi H, Sunami KS, et al. Impact of KRAS mutation on response and outcome of patients with stage III non-squamous non-small cell lung cancer. Cancer science 2015;106:1402–1407.

23. Hendriks LE, Kerr KM, Menis J, et al. Non-oncogene-addicted metastatic non-small-cell lung cancer: ESMO Clinical Practice Guideline for diagnosis, treatment and follow-up. Annals of oncology : official journal of the European Society for Medical Oncology 2023;34:358–376.

24. Hallqvist A, Enlund F, Andersson C, et al. Mutated KRAS Is an Independent Negative Prognostic Factor for Survival in NSCLC Stage III Disease Treated with High-Dose Radiotherapy. Lung cancer international 2012;2012:587424.

25. Riudavets M, Auclin E, Mosteiro M, et al. Durvalumab consolidation in patients with unresectable stage III non-small cell lung cancer with driver genomic alterations. Eur J Cancer 2022;167:142–148.

26. Mazieres J, Drilon A, Lusque A, et al. Immune checkpoint inhibitors for patients with advanced lung cancer and oncogenic driver alterations: results from the IMMUNOTARGET registry. Ann Oncol 2019;30:1321–1328.

27. Hirsch FR, Scagliotti GV, Mulshine JL, et al. Lung cancer: current therapies and new targeted treatments. Lancet 2017;389:299–311.

28. Liu Y, Zhang Z, Rinsurongkawong W, et al. Association of Driver Oncogene Variations With Outcomes in Patients With Locally Advanced Non-Small Cell Lung Cancer Treated With Chemoradiation and Consolidative Durvalumab. JAMA Netw Open 2022;5:e2215589.

29. Cortiula F, De Ruysscher D, Steens M, et al. Adjuvant durvalumab after concurrent chemoradiotherapy for patients with unresectable stage III NSCLC harbouring uncommon genomic alterations. Eur J Cancer 2023;184:172–178.

30. Skoulidis F, Li BT, Dy GK, et al. Sotorasib for Lung Cancers with KRAS p.G12C Mutation. The New England journal of medicine 2021;384:2371–2381.

31. Tsuboi M, Herbst RS, John T, et al. Overall Survival with Osimertinib in Resected EGFR-Mutated NSCLC. The New England journal of medicine 2023;389:137–147.

32. Solomon BJ, Ahn JS, Dziadziuszko R, et al. LBA2 ALINA: Efficacy and safety of adjuvant alectinib versus chemotherapy in patients with early-stage ALK+ non-small cell lung cancer (NSCLC). Annals of Oncology 2023;34:S1295–S1296 %@ 0923-7534.

