## Supplementary figures and images for "Equalizing prognostic disparities in stage III KRAS-mutant NSCLC: addition of durvalumab to combined chemoradiotherapy improves survival"

### Supp Figures 1-2

#### Eklund et al. Supplemental Figure 1

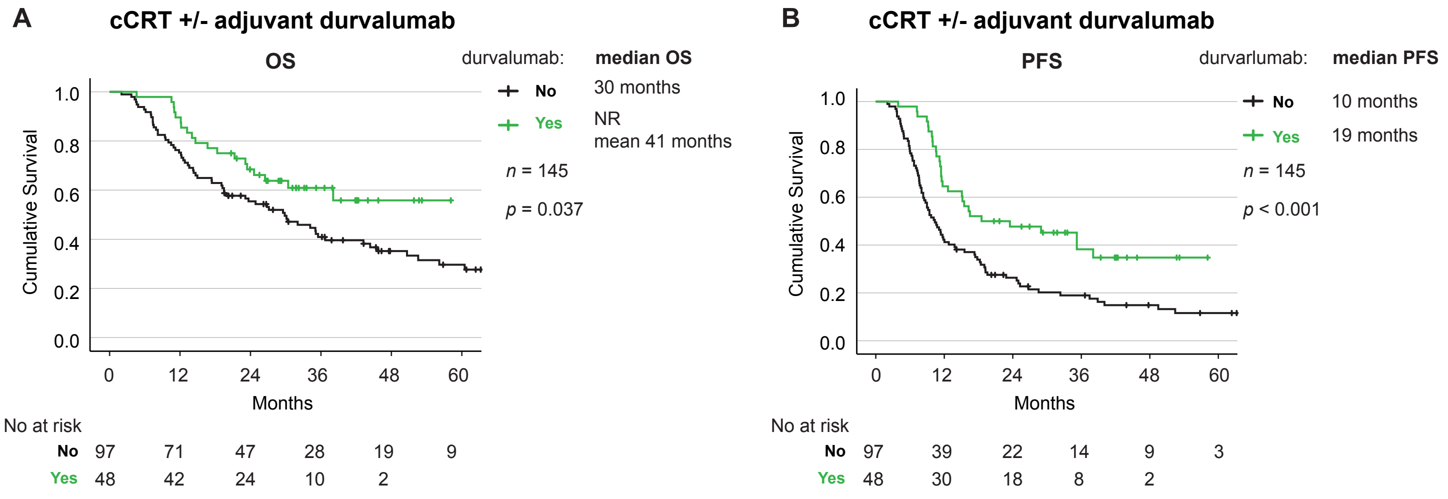

#### Eklund et al. Supplemental Figure 2

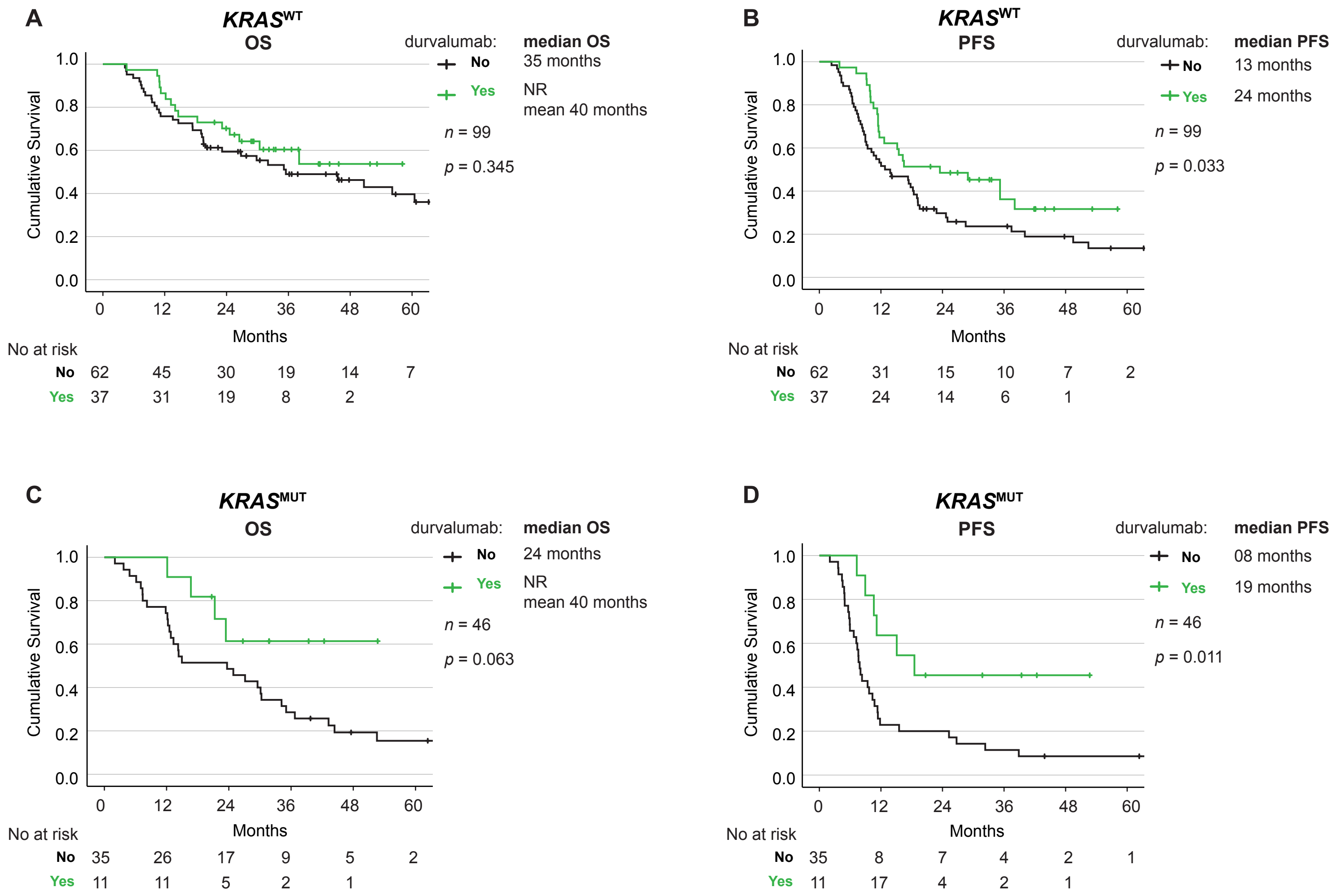
